# Nonlinear relationship between systolic blood pressure and diabetic retinopathy in type 2 diabetic patients in southern China: a single-center observational study

**DOI:** 10.1101/2022.10.18.22281231

**Authors:** Yisheng Luo, Chen Zhang, Chaochao Zhou, Zhisong Yan, Zezhong Ouyang, Wenbin Zhu

## Abstract

**Background:** Blood pressure (BP) control has been shown in clinical trials to reduce the risk of diabetic retinopathy (DR). To some extent, systolic blood pressure (SBP) has been shown to have a positive correlation with DR. However, there are no studies that have standardized SBP thresholds for DR prevention. Our goal was to use threshold analysis to further investigate the relationship between SBP and DR, identifying safe levels of SBP that contribute to DR prevention

**Methods:** We analyzed data from a cross-sectional study (December 2017-November 2018, n = 426, mean age 59.15±13.68) of patients with type 2 diabetes mellitus in the endocrinology department of Guangdong Provincial People’s Hospital, which is publicly available in the Dryad database. DR severity was assessed by retinal photographs. The International Clinical Diabetic Retinopathy and Diabetic Macular Edema Disease Severity Scale was used to classify DR severity and divide it into two groups: with DR and without DR. SBP was analyzed as a continuous variable. Multivariate logistic regression models, smoothed curve fitting, threshold analysis, and interaction tests were used to assess the relationship between SBP and DR.

**Results:** Prevalence of DR in the study population was 39.20%. After adjusting for age, sex, DM duration, HbA1c, BUN, HDL, LDL, TRIG, CHOL, TP, DBP, PP, eGFR, and CKD stage, the association between SBP and DR appears as a threshold effect, with a inflection point of 132 mm Hg. The risk of DR did not change significantly when SBP ≤132 mm Hg (OR: 0.86; 95% CI: 0.63 to 1.17, p=0.3400). When SBP ≥132 mm Hg, each 10 mm Hg increase in SBP raise the risk of developing DR by 28% (OR:1.28; 95% CI:1.07 to 1.54, P=0.0081). In addition, a stronger association of SBP with DR in patients with TP≤60g/L (OR=1.58, 95% CI: 1.19-2.08, P=0.001) compared to those with TP>60g/L (OR=1.15, 95% CI: 1.03, 1.27, P=0.012) was discovered, with a P value for interaction=0.023.

**Conclusion:** In Chinese patients with type 2 diabetes, SBP was significantly associated with DR when SBP ≥132 mm Hg. Further longitudinal studies are needed to confirm our findings, especially in patients with TP≤60g/L.

## Introduction

Diabetic retinopathy (DR) is a type of retinal microangiopathy caused by diabetes that is classified into three types: vascular proliferation, vascular leakage, and retinal ischemia. Diabetes is now recognized as a major cause of retinal damage and vision loss in diabetic patients, which is a major public health issue worldwide due to its very high and continuing prevalence. Diabetes is expected to affect 592 million people worldwide by 2035. Adult diabetes is prevalent in China at around 11.6%, while prediabetes is even more prevalent at 50.1% [1-3]. According to one study, approximately 93 million people worldwide have DR, 17 million of whom have proliferative diabetic retinopathy (PDR), and 10% are at risk for vision-threatening conditions. Blood pressure (BP), particularly systolic blood pressure (SBP), has been shown to be an important risk factor for DR in addition to other causes such as poor glycemic control [4-6]. In order to prevent cardiovascular complications such as coronary heart disease, guidelines for the prevention, detection, evaluation, and management of hypertension in adults in 2017 recommend a blood pressure goal of 130/80 mm Hg in adults with diabetes [7].

However, there are few research guidelines that standardize the target blood pressure values for DR prevention at this time. Although SBP has been shown to be positively correlated with DR within a certain range in previous studies, no specific SBP ranges or cut-off values have been detailed by threshold analysis [8, 9].

As a direct consequence, it is unclear whether the relationship between SBP and DR has a threshold or saturation effect. Given China’s large population base and the fact that evidence from Chinese patients is less than that from Western countries, as well as some methodological flaws in previous studies, we aimed to conduct a retrospective cross-sectional study to investigate the specific relationship between SBP components and the presence of DR in a southern Chinese population. We intended to refine the SBP cut-off values associated with DR using threshold analysis in order to better guide the range of SBP that should be controlled in diabetic patients to prevent DR.

## Methods

### Study population

The data for this study arrived from a publicly available database on the Internet, which was created and owned by the Department of Endocrinology at Guangdong Provincial People’s Hospital in China. The database contains health-care information on 503 type 2 diabetes patients in southern China, without information that could be used to identify individual participants. The data was collected between December 2017 and November 2018. There are 36 variables in the database that cover demographic, physical examination, and laboratory data information [10]. Please see the URL “https://datadryad.org/stash/dataset/doi:10.5061%2Fdryad.6kg1sd7“ and the reference for an additional detailed description of the database. By excluding patients with missing DR data, our study included 426 people.

This study is a retrospective cohort study. From December 2017 to November 2018, we collected data on 426 patients with type 2 diabetes who visited the endocrinology department of Guangdong Provincial People’s Hospital. The information on these patients was gathered in a non-selective and continuous manner. The following were the inclusion criteria: (1) patients with type 2 diabetes (according to World Health Organization criteria) [11]; (2) 35-degree 7-standard fields color retinal photographs from the Early Treatment Diabetic Retinopathy Study (ETDRS) [12]. The following were the exclusion criteria: (1) exclusion of any other ocular disease that may affect ocular circulation (e.g., glaucoma, endophthalmitis, retinal vascular obstruction, age-related macular degeneration, refractive error greater than 3 degrees, ocular trauma); (2) any serious systemic disease (e.g., myocardial infarction, cerebral infarction, connective tissue disorder); (3) previous history of intravitreal injection or dialysis; (4) pregnant or menstruating women. Because patient information was collected retrospectively and has been anonymized and de-traced, signed informed consent was not required.

### Assessment of DR

The presence or absence of DR was the primary outcome variable in this study. According to the International Clinical Diabetic Retinopathy and Diabetic Macular Edema Disease Severity Scale [13], the presence of DR was recorded as 1 while absence as 0.

### Measurement of BP

The target exposure variable was SBP, which was recorded as a continuous variable. The BP detection procedure was as follows: after five minutes of rest, an Omron electronic BP monitor was applied to the right arm of the seated participants. Two blood pressure (BP) measurements were taken five minutes apart. If the difference was greater than 10 mm for SBP and 5 mm for DBP, a third measurement was taken. The mean of the two closest readings was used as the BP value [14]. The difference between SBP and DBP (SBP-DBP) was used to calculate pulse pressure (PP).

### Covariates

The covariates included in this study were: sex, age, diabetes mellitus duration (DM Duration), glycated hemoglobin (HbA1c), urea nitrogen (BUN), high-density lipoprotein (HDL), low-density lipoprotein (LDL), triglycerides (TRIG), total cholesterol (CHOL), total protein (TP), diastolic blood pressure (DBP), pulse pressure (PP), glomerular filtration rate (eGFR), and chronic kidney disease stage (CKD stage). At the initial phase, all covariates were recorded. The rationale for inclusion was based primarily on previous work, our clinical experience, and previous literature that used DR as an outcome variable [9, 10, 15-17].

### Statistical analysis

Continuous variables were expressed as mean ± standard deviation (SD), while categorical variables were expressed as numbers and their percentages. We used multivariate multiple imputation, based on 5 replication and a chained equation approach method in the R MI procedure, to account for missing data. The missing values in each set included: HbA1c (n=7), BUN (n=1), HDL (n=4), LDL (n=4), TRIG (n=5), CHOL (n=5), and TP (n=1). After controlling for the following covariates that were suspected of being risk factors for DR, multivariate logistic regression models were built to assess the relationship between SBP and DR. Model I was adjusted for age and sex. Age, sex, DM Duration, HbA1c, BUN, HDL, LDL, TRIG, CHOL, TP, DBP, PP, eGFR, and CKD stage were all adjusted for in Model II. Smoothed curve fitting and threshold effect analysis were also used to address the potential nonlinear relationship between SBP and DR. Finally, subgroup analysis was performed to evaluate the following variables: age, sex, DM Duration, HbA1c, BUN, HDL, LDL, TRIG, CHOL, TP, DBP, PP, eGFR, CKD stage.

Statistical analyses were performed using the statistical package R software version 3.4.3 (https://www.R-project.org, R Foundation) and EmpowerStats version 4.1 (https://www.empowerstats.com; X&Y Solutions, Inc.). P-values < 0.05 were considered to be statistically significant.

## Results

### Descriptive analysis

Table 1 shows the patients’ baseline characteristics. According to the inclusion and exclusion criteria, 426 participants were included. We classified DR into two categories: With and Without DR groups. We noticed trends in the distribution of variables among the different subgroups after grouping. The patients’ average age was 59.15±13.68 years old. The prevalence of DR in patients was 39.20% (167/426). We found no statistically significant differences in the distribution of age, DBP, HbA1c, HDL, and TRIG among the various DR subgroups (all P values were greater than 0.05). The With DR group had a higher proportion of women, older age, higher SBP and PP, longer DM duration, higher BUN levels, higher LDL and CHOL levels, and higher CKD stage than the Without DR group. In contrast, we found lower TP and eGFR levels in the Without DR group when compared to the With DR group.

**Table 1.** Participants characteristics. (N=426)

| **DR** | **Without DR** | **With DR** | **P-value\*** |
| --- | --- | --- | --- |
| **N** | 259 | 167 |  |
| **Age** | 57.75 (14.16) | 60.41 (12.90) | 0.050 |
| **Sex** |  |  | 0.001 |
| **Male** | 162 (62.55%) | 78 (46.71%) |  |
| **Female** | 97 (37.45%) | 89 (53.29%) |  |
| **SBP (mm Hg)** | 135.21 (19.01) | 143.28 (23.31) | <0.001 |
| **SBP/10 (10 mm Hg)** | 13.52 (1.90) | 14.33 (2.33) | <0.001 |
| **DBP (mm Hg)** | 80.22 (11.60) | 80.15 (12.47) | 0.950 |
| **PP (mm Hg)** | 54.98 (16.09) | 63.13 (19.60) | <0.001 |
| **DM duration (years)** | 6.00 (1.00-59.00) | 10.00 (1.00-31.00) | <0.001 |
| **HbA1c (%)** | 9.73 (2.39) | 9.71 (2.28) | 0.937 |
| **BUN** | 5.37 (2.06-19.86) | 6.13 (2.40-39.60) | <0.001 |
| **HDL (mmol/L)** | 1.00 (0.33) | 1.07 (0.32) | 0.051 |
| **LDL (mmol/L)** | 3.12 (0.91) | 3.37 (1.11) | 0.010 |
| **TRIG (mmol/L)** | 1.52 (0.44-32.85) | 1.51 (0.52-20.12) | 0.989 |
| **CHOL (mmol/L)** | 4.87 (1.39) | 5.27 (1.85) | 0.012 |
| **TP (g/L)** | 65.68 (5.72) | 64.46 (6.53) | 0.044 |
| **eGFR (mL/min/1.73 m<sup>2</sup>)** | 92.58 (23.95-294.98) | 80.29 (7.24-374.89) | <0.001 |
| **CKD stage** |  |  | <0.001 |
| **1** | 141 (54.44%) | 63 (37.72%) |  |
| **2** | 89 (34.36%) | 52 (31.14%) |  |
| **3** | 24 (9.27%) | 29 (17.37%) |  |
| **4** | 5 (1.93%) | 14 (8.38%) |  |
| **5** | 0 (0.00%) | 9 (5.39%) |  |

Abbreviations: DR: diabetic retinopathy; SBP: systolic blood pressure; SBP/10: systolic blood pressure divided by 10; DBP: diastolic blood pressure; PP: pulse pressure; DM: diabetes mellitus; HbA1c: glycated hemoglobin; BUN: blood urea nitrogen; HDL: high-density lipoprotein; LDL: low-density lipoprotein; TRIG: triglyceride; CHOL: total cholesterol; TP: total protein; eGFR: estimated glomerular filtration rate; CKD: chronic kidney disease; SD, standard deviation.
\*P value: Kruskal Wallis Rank Test for continuous variables, Fisher Exact for categorical variables with Expects<10.
Data presented are Mean (SD) /Median (Min-Max) / n (%)

### Linear relationship between SBP and DR

Table 2 shows the results of different covariate adjustment strategies used to examine the relationship between SBP and DR. Each 10 mm Hg increase in SBP was associated with a 20% increase in the risk of DR in the unadjusted model (OR: 1.20, 95% CI: 1.09 to 1.32). After adjusting for demographic factors, every 10 mm Hg increase in SBP increased the risk of DR by 18% (OR: 1.18, 95% CI: 1.07 to 1.31). After controlling for Age, Sex, DM duration, HbA1c, BUN, HDL, LDL, TRIG, CHOL, TP, DBP, PP, eGFR, and CKD stage, no significant association was found between SBP and DR (OR: 1.15, 95% CI: 0.99 to 1.33). We transformed SBP into a categorical variable based on quadratic grouping for the purpose of sensitivity analysis and calculated the p-value for the trend test. The findings revealed that when SBP was used as a continuous variable, the results were inconsistent with those as a categorical variable.

**Table 2.** Relationship between SBP and DR in different models.

| Per 10 mm Hg<br>increased in<br>SBP | unadjusted model | Adjust I model | Adjust II model |
| --- | --- | --- | --- |
|  | (OR, 95%CI, P) | (OR, 95%CI, P) | (OR, 95%CI, P) |
| **SBP/10** | 1.20 (1.09, 1.32) <0.001 | 1.18 (1.07, 1.31) <0.001 | 1.15 (0.99, 1.33) 0.063 |
| **SBP/10** |  |  |  |
| **Quartiles** |  |  |  |
| **Q1 (8.0-12.3)** | Ref | Ref | Ref |
| **Q2 (12.4-13.5)** | 0.73 (0.40, 1.32) 0.295 | 0.71 (0.39, 1.30) 0.269 | 0.44 (0.18, 1.10) 0.079 |
| **Q3 (13.6-15.2)** | 1.06 (0.61, 1.85) 0.837 | 0.93 (0.52, 1.68) 0.809 | 0.40 (0.10,1.52) 0.176 |
| **Q4 (15.3-21.9)** | 2.16 (1.23, 3.79) 0.007 | 1.99 (1.11, 3.58) 0.021 | 0.38 (0.03,4.20) 0.428 |

Unadjusted model: did not adjust any covariant.
Adjusted I model: adjusted age and sex.
Adjust II model: adjusted age, sex, DM duration, HbA1c, PP, BUN, HDL, LDL,
TRIG, CHOL, TP, eGFR and CKD stage.
Abbreviations: SBP/10: systolic blood pressure divided by 10; OR: odds ratio; CI:
confidence interval; Ref: reference.

### Non-linear association of SBP with DR

We discovered a nonlinear relationship between SBP and DR using smoothed curve fitting and a generalized summation model. After adjusting for age, sex, DM duration, HbA1c, BUN, HDL, LDL, TRIG, CHOL, TP, DBP, PP, eGFR, and CKD stage, the findings (Fig 1) show that the association between SBP and DR appears as a threshold effect. We calculated the SBP inflection point to be 132 mm Hg using a two-piecewise linear model and a recursive algorithm. The risk of DR did not change significantly with increasing SBP on the left side of the inflection point (OR: 0.86; 95% CI: 0.63 to 1.17, p=0.3400). Each 10 mm Hg increase in SBP to the right of the inflection point increased the risk of developing DR by 28% (OR:1.28; 95% CI:1.07 to 1.54, P=0.0081) (Table 3).

**Table 3.**
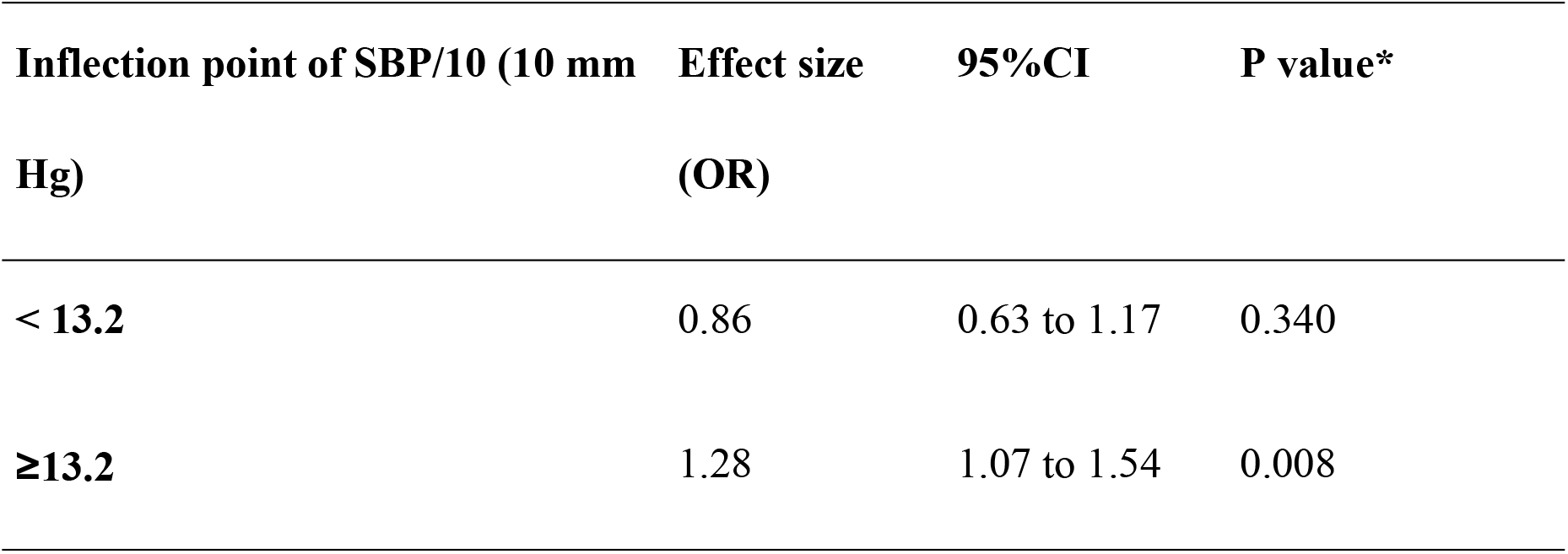
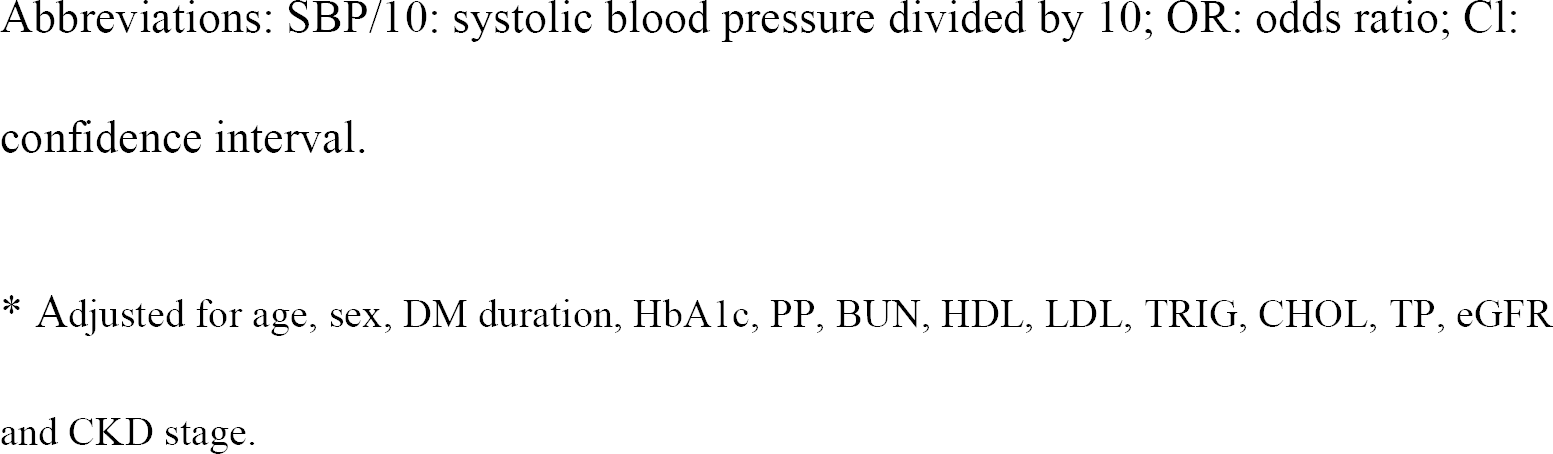
Threshold Effect Analysis of SBP and DR using Piece-wise Linear Regression.

| Inflection point of SBP/10 (10 mm Hg) | Effect size (OR) | 95%CI | P value* |
| --- | --- | --- | --- |
| < 13.2 | 0.86 | 0.63 to 1.17 | 0.340 |
| ≥13.2 | 1.28 | 1.07 to 1.54 | 0.008 |

Abbreviations: SBP/10: systolic blood pressure divided by 10; OR: odds ratio; CI: confidence interval.
\* Adjusted for age, sex, DM duration, HbA1c, PP, BUN, HDL, LDL, TRIG, CHOL, TP, eGFR and CKD stage.

**Fig 1.**
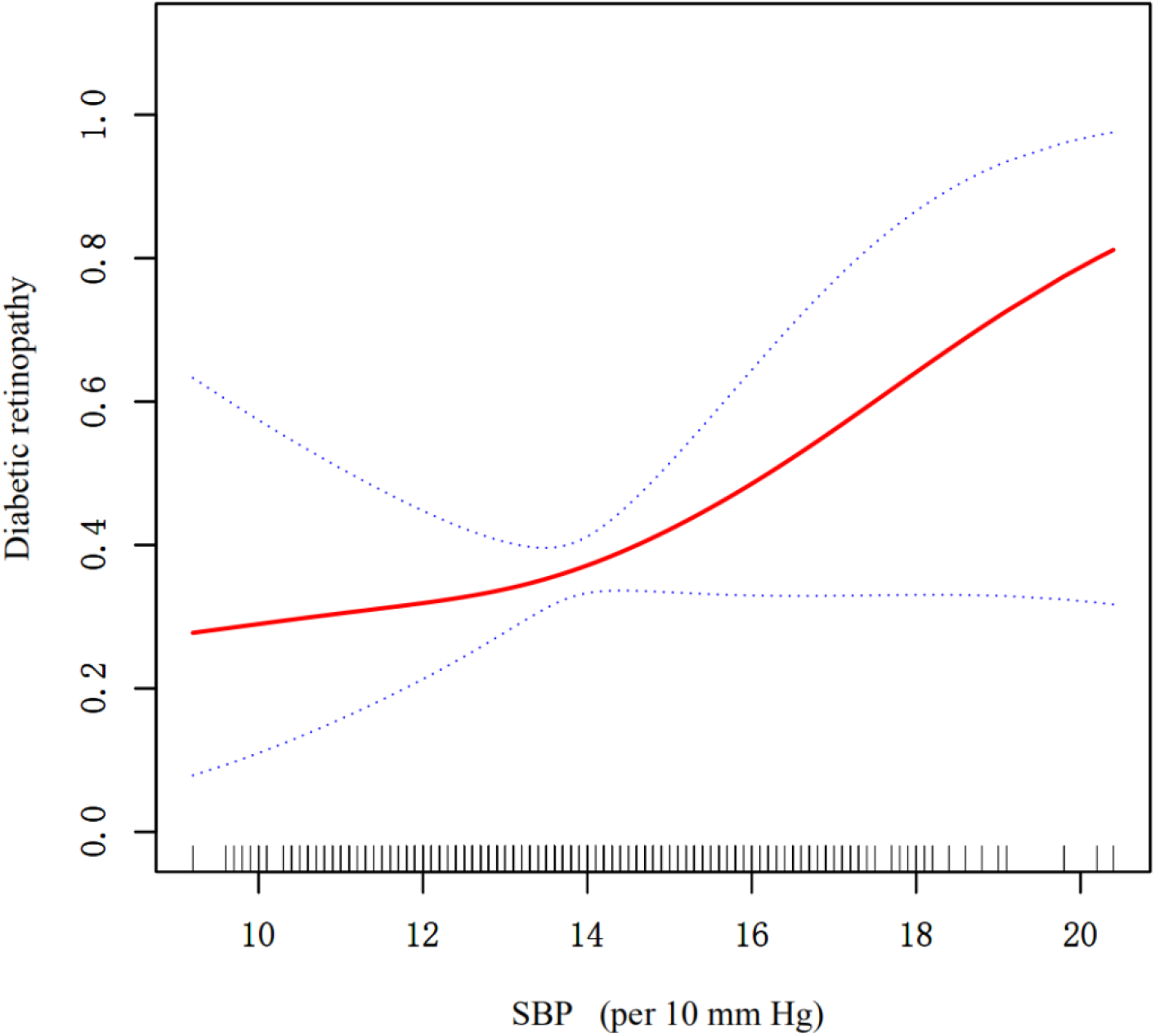
Association between systolic blood pressure (SBP) and diabetic retinopathy (DR). A threshold, nonlinear association between SBP and DR was found in a generalized additive model (GAM). Solid rad line represents the smooth curve fit between variables. Blue bands represent the 95% of confidence interval from the fit. All adjusted for age, sex, DM duration, HbA1c, BUN, HDL, LDL, TRIG, CHOL, TP, DBP, PP, eGFR, CKD stage.

### Analysis of the relationship between SBP and DR in different prespecified subgroups

We included age (≤60 years/>60 years), sex(Male/Female), DM duration (≤10 years/>10 years), HbA1c (≤8%/>8%), DBP (≤80 mm Hg/>80 mm Hg), PP (21.00-48.00 mm Hg/49.00-63.00 mm Hg/64.00-125.00 mm Hg), BUN (≤7.1 mmol/L/>7.1 mmol/L), HDL (≤0.9 mmol/L/>0.9 mmol/L), LDL (≤3.4 mmol/L, >3.4 mmol/L), TRIG (≤1.69 mmol/L/>1.69 mmol/L), CHOL (≤5.72mmol/L/>5.72 mmol/L), TP (≤ 60g/L/>60 g/L), eGFR (≥90mL/min/1.73 m^2/≥60 and <90 mL/min/1.73 m^2/<60mL/min/1.73 m^2) as the pre-defined effect modifiers to explore whether the association between SBP and DR was stable across subgroups. Fig 2 showed that when age, sex, DM duration, HbA1c, PP, BUN, HDL, LDL, TRIG, CHOL, eGFR were used as potential effect modifiers, the association of SBP with DR was not observed to be significantly different between subgroups (P for interaction >0.05). In addition, we observed a stronger association of SBP with DR in patients with TP≤ 60g/L (OR=1.58, 95% CI: 1.19-2.08, P=0.001) compared to those with TP>60g/L (OR=1.15, 95% CI: 1.03, 1.27, P=0.012), with a P value for interaction=0.023.

**Fig 2.**
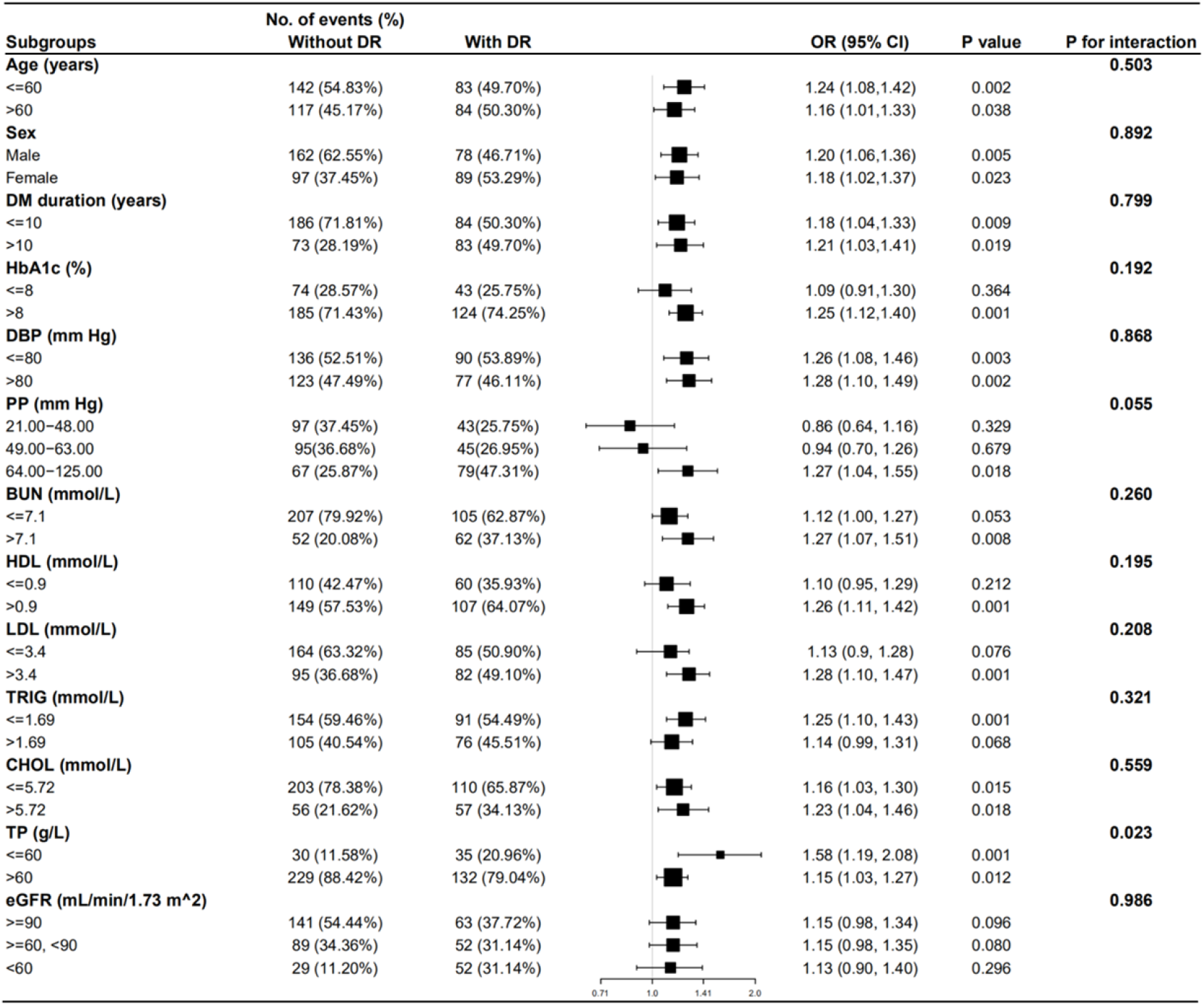
Effect size of SBP on DR in prespecified and exploratory subgroups in Each Subgroup. Adjusted for age, sex, DM duration, HbA1c, BUN, HDL, LDL, TRIG, CHOL, TP, DBP, PP, eGFR.

## Discussion

The association between SBP and DR was investigated in this retrospective cohort study. We discovered a threshold effect relationship between SBP and DR by analyzing the data of 426 patients. The findings revealed that when SBP exceeded 132 mm Hg, the risk of DR increased by 28% for every 10 mm Hg increase in SBP (OR:1.28; 95% CI:1.07 to 1.54, P=0.0081). The effect of SBP on DR was more pronounced in the TP ≤60 population (OR=1.58, 95% CI: 1.19-2.08, P=0.001). However, due to the small number of patients in our study with TP ≤60, the effect of SBP on DR in this group could not be accurately elaborated, which will be one of the priority groups for future studies on the relationship between SBP and DR.

Our results are the same as those of Liu L, et al. and Barak Rosenn, et al. The former observed a positive association between SBP and DR (OR : 1.45, 95% CI: 1.28 to 1.65) in a cross-sectional study involving 2189 Singaporean diabetic patients aged 40-80 years old [9]. While Barak Rosenn, et al. found higher blood pressure was associated with DR in a cohort study of 154 US women with diabetes (OR: 2.1, 95% CI: 1.7 to 3.6) [17]. These findings have some similarities with our conclusions. However, in the fully adjusted model (Adjust II model), we discovered that the relationship between SBP and DR was not linear (supplementary information Fig 1). We hypothesize that some residual confounding factors may obscure the true relationship between SBP and DR. The level of TP, for example, may affect the patient’s SBP, whereas LDL, TRIG, DBP, PP, eGFR, and CKD stage may influence the development of DR in diabetes patients [8-10, 15, 17, 18]. As a result, we used more outcome adjusted covariates in the Chinese population than in the previous researches.

The pathophysiology of the relationship between SBP and DR has not been fully elucidated. However, it has been pointed out that normal retinal autoregulation can keep blood flow constant in retinal blood vessels under different perfusion pressures, preventing endothelial cells from long-term exposure to high levels of glucose and other factors, which avoid the retina from systemic hypertension damage. When diabetes is combined with hypertension, retinal blood flow increases and accelerates the development and progression of retinal lesions [18]. In our study, in addition to confirming the positive association between SBP and DR within a certain range, the threshold effect analysis was used to identify the fold point, which suggested that for every 10 mm Hg increase in SBP when SBP>132 mm Hg, the risk of DR increased by 28% compared to patients with SBP≤132 mm Hg. The findings of this study can help diabetic patients reduce their risk of DR by controlling SBP, filling a gap in previous research. Furthermore, we went on to identify a subpopulation with TP≤60, where the effect of SBP on DR was more significant. However, due to the small number of patients with TP≤60 in the research, the effect of SBP on DR in this population could not be accurately elaborated, which will be one of the important directions for future research into the relationship between SBP and DR.

The main strengths of this study were as follows: (1) the relatively large sample size. (N=426) (2) we used multivariate multiple imputation, based on 5 replication and a chained equation approach method in the R MI procedure, to account for missing data. The missing values in each set included: HbA1c (n=7), BUN (n=1), HDL (n=4), LDL (n=4), TRIG (n=5), CHOL (n=5), and TP (n=1). (3) Compared to previous studies, the current study adjusted for more covariates, ensuring the results more stable. Except for the basic population characteristics, Liu L, et al. only adjusted for diabetes duration, antidiabetic drug use, total cholesterol, HDL, glycated hemoglobin, antihypertensive drug use, CVD history and CKD status. Based on previous literature reports and considering the result of, we appropriately added BUN, LDL, TRIG,TP, DBP, PP, eGFR, and CKD stage as covariates. According to previous literature, we added BUN, LDL, TRIG, TP, DBP, PP, eGFR, and CKD stage as covariates, and the results were more stable and reliable. (4) In this study, we used sensitivity analysis and an algorithm to reveal the nonlinearity, which more accurately reflects the true relationship between SBP and DR [8-10, 15, 17, 18].

There are several drawbacks of this study. Pregnant, menstruating patients were excluded from this study. Therefore, future use of the findings of this study in the population of pregnant or menstruating women requires caution. Observational study is inevitably confounded by confounding factors, though we rigorously adjusted for confounding factors and assessed the robustness of the results by sensitivity analysis. Limited to the nature of observational studies, we could only observe associations instead of assessing causality. We could only adjust for measurable confounding but not for unmeasurable confounding, so future clinical studies with higher levels of evidence in larger populations to validate our findings are warranted.

## Conclusions

In conclusion, our study demonstrated that SBP≥132 mm Hg was significantly associated with the risk of DR in patients with type 2 diabetes in southern China (OR between SBP/10 and DR: 1.28, 95% CI: 1.07 to 1.54), which suggests that to prevent the risk of DR in diabetic patients, their SBP should be controlled at <132 mm Hg. Further analysis revealed a distinct population with TP≤60. The association between SBP and DR was stronger in patients with TP≤60g/L (OR=1.58, 95% CI: 1.19-2.08, P=0.001) compared to those with TP>60g/L (OR=1.15, 95% CI: 1.03, 1.27, P=0.012), with an interaction P=0.023. This will also be one of the important directions for future studies on the relationship between SBP and DR.

## Data Availability

All data files are available from the Dryad database (URL "https://datadryad.org/stash/dataset/doi:10.5061%2Fdryad.6kg1sd7").

https://datadryad.org/stash/dataset/doi:10.5061%2Fdryad.6kg1sd7

## Acknowledgements

The authors acknowledge Dryad database for providing their platforms and contributors from the Department of Endocrinology of Guangdong Provincial People’s Hospital for uploading their meaningful datasets to make this and other studies possible.

## Author contributions

**Conceptualization:** Yisheng Luo.

**Data Curation:** Yisheng Luo, Chen Zhang.

**Formal Analysis:** Yisheng Luo, Chaochao Zhou, Zhisong Yan, Zezhong Ouyang.

**Methodology:** Yisheng Luo, Zhisong Yan, Zezhong Ouyang, Wenbin Zhu.

**Supervision:** Yisheng Luo, Chen Zhang.

**Writing – Original Draft:** Yisheng Luo, Zhisong Yan, Zezhong Ouyang, Wenbin Zhu.

**Writing – Review & Editing:** Yisheng Luo, Chen Zhang, Chaochao Zhou.

